# The Role of Health Professionals in Managing Type 2 Diabetes Mellitus in Community: A Scoping Review

**DOI:** 10.1101/2024.11.28.24318129

**Authors:** Yourisna Pasambo, Moses Glorino Rumambo Pandin, Ferry Efendi

## Abstract

**Background:** Diabetes Mellitus Type 2 (T2DM) is a global health problem that continues to increase. Suboptimal treatment can worsen the patient’s condition, cause more serious complications, and increase the death rate. The role of various health professionals in the management of T2DM in the community is increasingly recognized as an important element for achieving better outcomes. Objective of this review is to identify and analyze the role of each health professional and their challenges in treating T2DM.

**Method:** Review of the scope of the main research that discusses it the role of the role of each health professional health professional in treating T2DM in the community. Various databases were searched via Web of Science, Scopus, PubMed, and ProQuest. Four databases were searched and 462 articles were retrieved. After screening articles and abstracts, 51 full texts were assessed for eligibility, and finally 9 studies were further analyzed and synthesized.

**Results:** seven quantitative studies and two qualitative studies met the inclusion criteria. Four themes emerged: (1) the role of each health professional, (2) Determining factors for the success of T2DM treatment, (3) Challenges in treating T2DM, and (4) Health policy and support systems.

**Conclusion:** found unclear role definitions for healthcare professionals managing T2DM, leading to overlapping responsibilities and inefficiencies in patient care. This study highlights the importance of a clear and standardized treatment protocol and guideline, containing clear roles for each health professional for the management of people with T2DM in the community. Governments need to promote IPC and provide the necessary resources and infrastructure to support collaborative practice resulting in better outcomes for T2DM patients.

## Introduction

Diabetes Mellitus Type 2 (T2DM) is a global health problem that continues to increase, with increasingly high prevalence in various countries, including Indonesia (World Health Organization, 2022). T2DM is a chronic disease that can cause serious complications, such as cardiovascular disease, blindness, kidney failure, and amputation (CDC, 2020). The increasing prevalence of T2DM is influenced by various factors, including unhealthy eating patterns, lack of physical activity, obesity, and genetic factors (Powers et al., 2015). According to data from the International Diabetes Federation (IDF), in 2021, it is estimated that 537 million adults in the world suffer from diabetes, and this figure is predicted to increase to 783 million in 2045 (IDF, 2021). In Indonesia, the prevalence of T2DM reaches 8.5% of the total adult population, which shows a very significant figure in the context of the public health burden (Kemenkes, 2021).

T2DM disease not only affects individuals, but also has a major impact on the country’s health and economic systems (Zhang et al., 2010). Complications caused by T2DM require long-term treatment costs and increase the burden on the health system (Wu et al., 2018). Suboptimal treatment of T2DM can worsen the patient’s condition, cause more serious complications, and increase the death rate (Karatzanis et al., 2013). In Indonesia, even though various diabetes control efforts have been made, the high prevalence of T2DM and the increasing number of complications indicate that the challenges in treating this disease are still large (Cholil et al., 2019).

The role of various health professionals in the management of T2DM in the community is increasingly recognized as an important element to achieve better outcomes in the management of this disease (McGill et al., 2017). A multidisciplinary approach involving doctors, nurses, nutritionists, physiotherapists, psychologists and other health professionals has been proven to be more effective in treating patients’ conditions holistically (Liu et al., 2021). However, although there is growing awareness of the importance of synergy within health teams, many challenges are faced in its implementation (Desse et al., 2023). Barriers to communication, coordination, and a lack of shared understanding of the role of each professional often hinder the effectiveness of managing T2DM in the community (Alshowair et al., 2023). In addition, limited resources and training for health professionals in implementing a multidisciplinary team approach are also significant obstacles (Riordan et al., 2023). There are no consistent standards for diabetes education and self-management strategies among health care providers. These inconsistencies can result in varying approaches to patient care, contributing to overlapping incidents and unclear roles (Dudley et al., 2014).

Strengthening synergy between health professionals in treating T2DM in the community requires a more structured and collaborative approach (Gilbert et al., 2010). A team-based approach involving multiple disciplines can improve coordination in patient care, improve health education, and help patients manage their conditions more effectively (Nurchis et al., 2022). One solution that has been proven effective is the use of a multidisciplinary team-based diabetes care management model, which involves doctors, nurses, nutritionists, and other experts to provide comprehensive care (American Diabetes Association, 2018). In addition, more intensive training for health professionals on the importance of teamwork and strengthening communication between professionals is also needed to achieve better results (Harcke et al., 2023). Strengthening health policies that support team approaches and community empowerment in the management of T2DM is also an important factor in strengthening this synergy (Black et al., 2017).

Comprehensive treatment of people with T2DM in the community requires the role and collaboration of all health professionals in the community. Understanding the role that has been carried out so far by health professionals is necessary to analyze the causes of inadequate treatment of T2DM in the community. This scoping review aims to provide additional knowledge from the perspective of the role of health professionals and the challenges faced in treating T2DM in the community. Therefore, this study aims to obtain a broad overview by exploring themes emerging from the literature, regarding the role of each health professional involved in treating T2DM in the community and the challenges faced.

## Methods

This scoping review was carried out based on the scoping review guidelines developed by (Arksey & O’Malley, 2005) and (Levac et al., 2010), carried out in five stages, namely identifying research questions, identifying relevant studies and selecting studies, mapping data, compile and summarize, and report results. The primary goal of conducting a scoping review is to map the available empirical evidence on a topic of interest and identify gaps in the literature. The strength of the scoping review methodology lies in its ability to give meaning and significance to the subject of interest and extract the essence from a diverse body of evidence. This scoping review was used to examine the scope, reach, and nature of relevant research (Davis et al., 2009). Traceability reporting (Figure 1) follows PRISMA recommendations (Page et al., 2021).

**Figure 1.**
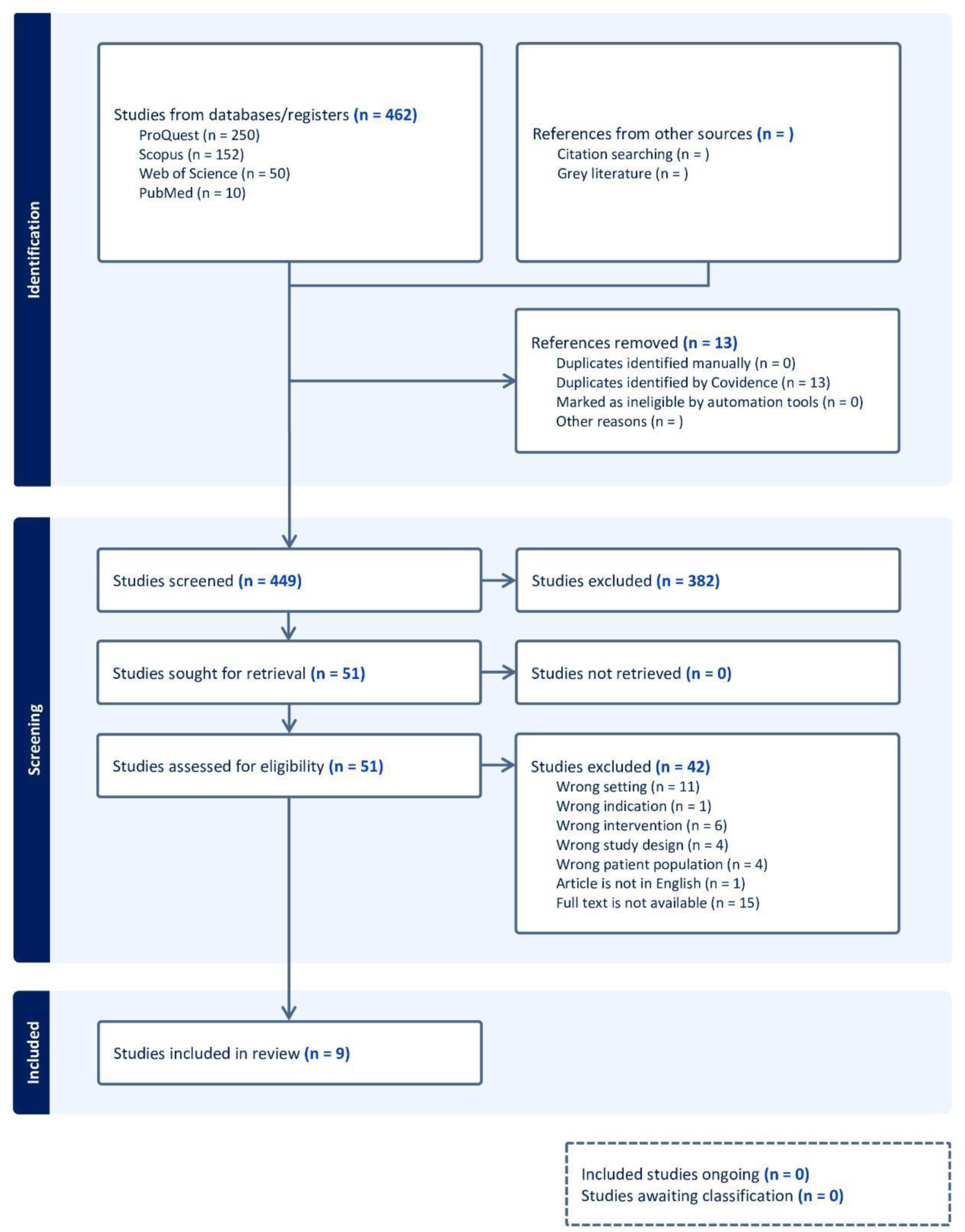
PRISMA flow diagram for review: the role of health professionals in the management of T2DM in the community

### Literature Search

We began our literature search by asking the question: What is the role of each health professional and what challenges are faced in treating T2DM in the community? The search was carried out using four scientific databases: PubMed, Scopus, Web of Science, ProQuest. The same key terms were used in the scientific databases mentioned above following the search query: (“healthcare professionals” OR “healthcare providers” OR doctors OR nurses OR dietitians OR pharmacists OR health educators OR physiotherapists) AND (“Diabetes Mellitus Type 2” OR "T2DM") AND (collaboration OR synergy OR "interprofessional cooperation") AND ("community health" OR "public health"). Searches were limited to language (English language publications) and title/abstract levels. According to these established criteria, 462 studies were found.

### Data retrieval

Inclusion criteria were as follows: research (a) was an empirical article, (b) published in a peer reviewed journal, (c) written in English, (d) focused on a topic of interest, and (e) involved a community-based T2DM population. or without comorbidities. Exclusion criteria were as follows: studies (a) were editorials, case writing, protocols, and reviews, and (b) involved T2DM patients other than those in the community. Data was taken systematically in two stages.

Our search yielded 462 publications that were uploaded to Covidence for evaluation. Covidence is an online software platform used to manage literature reviews by research teams (Covidence, 2024). There were 13 duplicate articles removed by the system and an additional 382 studies removed during title and abstract screening due to irrelevance. Of the 51 full articles reviewed, 42 studies were eliminated by the consensus team. The total number of studies included in the extract was 9 articles.

Two reviewers independently reviewed each publication using the template described below. Not all fields apply to all publications reviewed. At each stage of the review process, two reviewers independently review and make judgments. Covidence indicated any discrepancies in assessment and a third author reviewed with the initial two authors who completed the review. The selection of publications can be seen in the PRISMA flow diagram in Figure 1.

### Data extraction and analysis

To standardize data extraction procedures, made template in Covidence. Table 1 displays data extraction from 9 studies containing the following information: author, year, country, study aims, study design, participants, sample size, health professionals involved, and limitations. The extracted data was exported from Covidence into a Microsoft Excel spreadsheet for further analysis to explore answers to the stated research questions using qualitative content analysis according to the methodology (Mayring, 2014). The result of inductive theme development is a logical categorization of information from the included research based on the principles of thematic analysis. The methodological process consists of eight steps, including the formulation of research questions and description of theoretical background, definition of themes used as selection criteria for determining relevant material from the subject of analysis, and text coding including formulation of themes and assigned texts of the research. Furthermore, analysis focused on category/theme revision and comparison with research questions, final coding, development of main themes, and summative and narrative presentation of results.

**Table 1.**
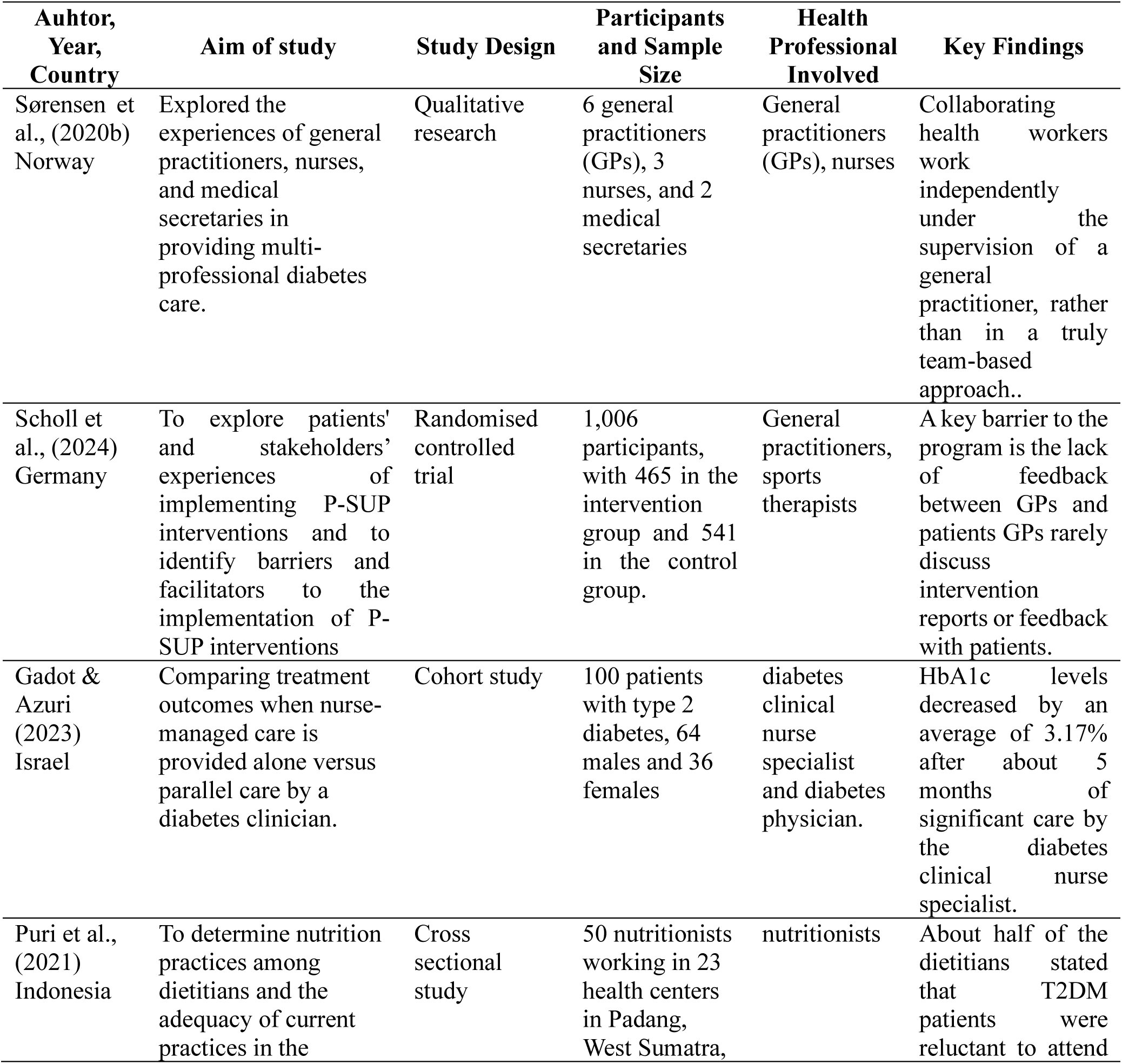
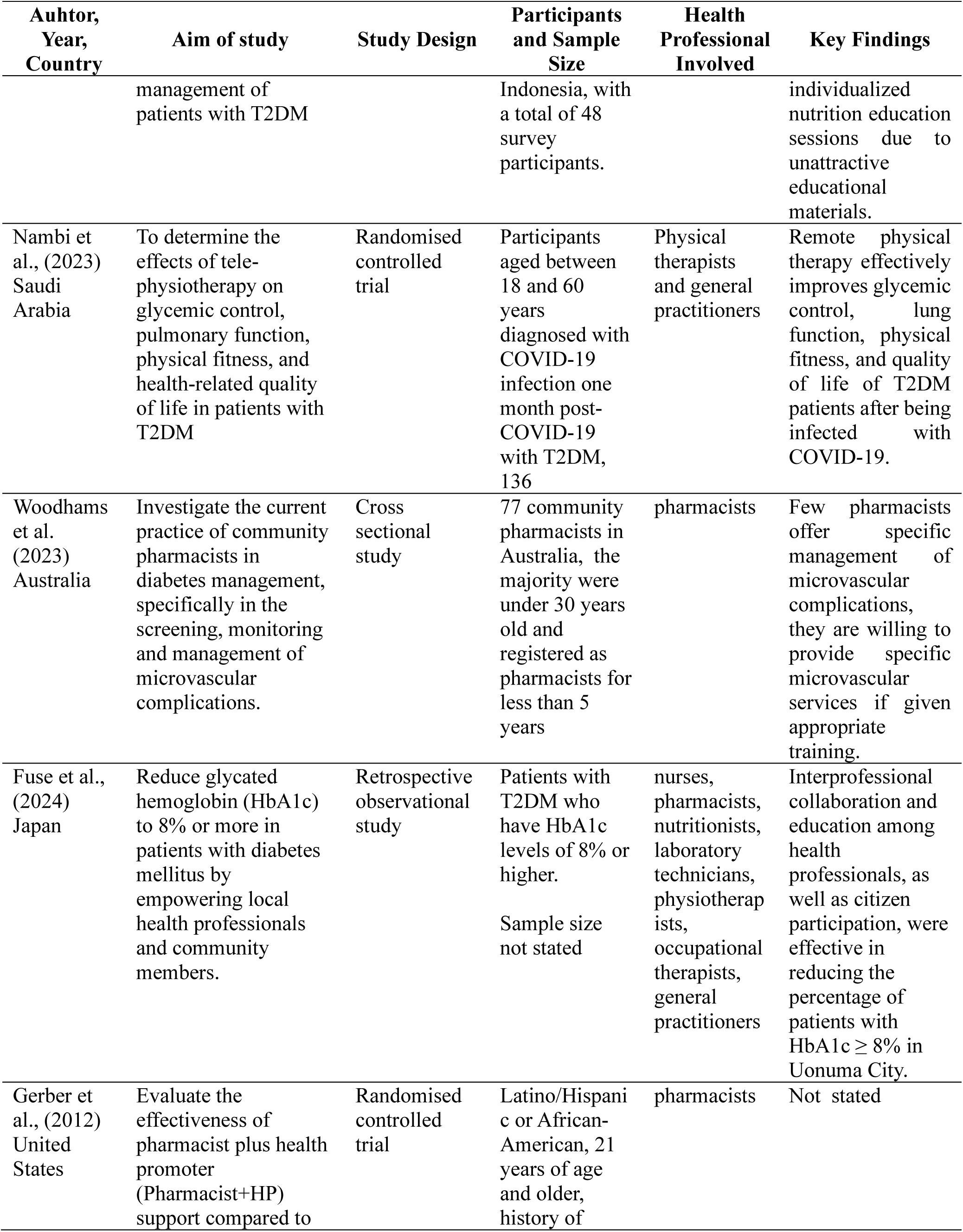
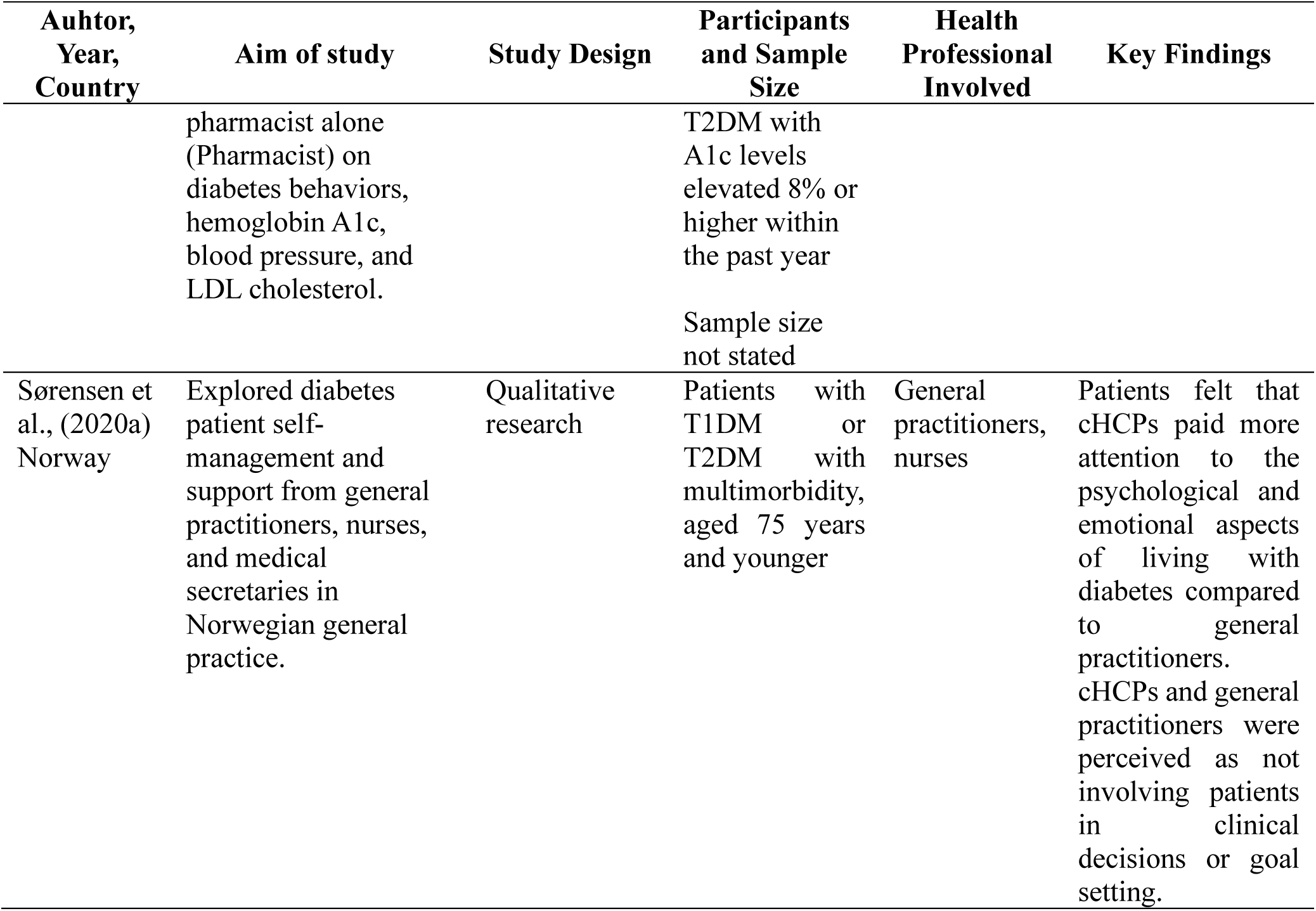
Summary of Included Studies.

| Auhtor,<br>Year,<br>Country | Aim of study | Study Design | Participants<br>and Sample<br>Size | Health<br>Professional<br>Involved | Key Findings |
| --- | --- | --- | --- | --- | --- |
| Sørensen et al., (2020b)<br>Norway | Explored the experiences of general practitioners, nurses, and medical secretaries in providing multi-professional diabetes care. | Qualitative research | 6 general practitioners (GPs), 3 nurses, and 2 medical secretaries | General practitioners (GPs), nurses | Collaborating health workers work independently under the supervision of a general practitioner, rather than in a truly team-based approach.. |
| Scholl et al., (2024)<br>Germany | To explore patients' and stakeholders' experiences of implementing P-SUP interventions and to identify barriers and facilitators to the implementation of P-SUP interventions | Randomised controlled trial | 1,006 participants, with 465 in the intervention group and 541 in the control group. | General practitioners, sports therapists | A key barrier to the program is the lack of feedback between GPs and patients GPs rarely discuss intervention reports or feedback with patients. |
| Gadot & Azuri (2023)<br>Israel | Comparing treatment outcomes when nurse-managed care is provided alone versus parallel care by a diabetes clinician. | Cohort study | 100 patients with type 2 diabetes, 64 males and 36 females | diabetes clinical nurse specialist and diabetes physician. | HbA1c levels decreased by an average of 3.17% after about 5 months of significant care by the diabetes clinical nurse specialist. |
| Puri et al., (2021)<br>Indonesia | To determine nutrition practices among dietitians and the adequacy of current practices in the | Cross sectional study | 50 nutritionists working in 23 health centers in Padang, West Sumatra, | nutritionists | About half of the dietitians stated that T2DM patients were reluctant to attend |

| **Auhtor,**<br>**Year,**<br>**Country** | **Aim of study** | **Study Design** | **Participants**<br>**and Sample**<br>**Size** | **Health**<br>**Professional**<br>**Involved** | **Key Findings** |
| --- | --- | --- | --- | --- | --- |
|  | management of patients with T2DM |  | Indonesia, with a total of 48 survey participants. |  | individualized nutrition education sessions due to unattractive educational materials. |
| Nambi et al., (2023)<br>Saudi Arabia | To determine the effects of tele-physiotherapy on glycemic control, pulmonary function, physical fitness, and health-related quality of life in patients with T2DM | Randomised controlled trial | Participants aged between 18 and 60 years diagnosed with COVID-19 infection one month post-COVID-19 with T2DM, 136 | Physical therapists and general practitioners | Remote physical therapy effectively improves glycemic control, lung function, physical fitness, and quality of life of T2DM patients after being infected with COVID-19. |
| Woodhams et al. (2023)<br>Australia | Investigate the current practice of community pharmacists in diabetes management, specifically in the screening, monitoring and management of microvascular complications. | Cross sectional study | 77 community pharmacists in Australia, the majority were under 30 years old and registered as pharmacists for less than 5 years | pharmacists | Few pharmacists offer specific management of microvascular complications, they are willing to provide specific microvascular services if given appropriate training. |
| Fuse et al., (2024)<br>Japan | Reduce glycated hemoglobin (HbA1c) to 8% or more in patients with diabetes mellitus by empowering local health professionals and community members. | Retrospective observational study | Patients with T2DM who have HbA1c levels of 8% or higher.<br>Sample size not stated | nurses, pharmacists, nutritionists, laboratory technicians, physiotherapists, occupational therapists, general practitioners | Interprofessional collaboration and education among health professionals, as well as citizen participation, were effective in reducing the percentage of patients with HbA1c ≥ 8% in Uonuma City. |
| Gerber et al., (2012)<br>United States | Evaluate the effectiveness of pharmacist plus health promoter (Pharmacist+HP) support compared to | Randomised controlled trial | Latino/Hispanic or African-American, 21 years of age and older, history of | pharmacists | Not stated |
|  | pharmacist alone<br>(Pharmacist) on<br>diabetes behaviors,<br>hemoglobin A1c,<br>blood pressure, and<br>LDL cholesterol. |  | T2DM with<br>A1c levels<br>elevated 8% or<br>higher within<br>the past year<br>Sample size<br>not stated |  |  |
| Sørensen et<br>al., (2020a)<br>Norway | Explored diabetes<br>patient self-management and<br>support from general<br>practitioners, nurses,<br>and medical<br>secretaries in<br>Norwegian general<br>practice. | Qualitative<br>research | Patients with<br>T1DM or<br>T2DM with<br>multimorbidity,<br>aged 75 years<br>and younger | General<br>practitioners,<br>nurses | Patients felt that<br>cHCPs paid more<br>attention to the<br>psychological and<br>emotional aspects<br>of living with<br>diabetes compared<br>to general<br>practitioners.<br>cHCPs and general<br>practitioners were<br>perceived as not<br>involving patients<br>in clinical<br>decisions or goal<br>setting. |

## Findings

A total of nine articles were included: seven quantitative and two qualitative. Studies were conducted in eight countries, two studies in Norway, and one study each in Saudi Arabia, Australia, Germany, Israel, Indonesia, Japan, and the United States. The majority of studies involved doctors and nurses in the management of T2DM in the community. Some studies involve other health professions such as pharmacists, nutritionists, health promotion workers, physiotherapists, psychologists, physiotherapists, and even sports therapists. Results from all articles reflect experiences of health professionals’ roles and perceived barriers during treatment of T2DM in the community (Table 1), while themes identified are as depicted in Table 2.

**Table 2.**
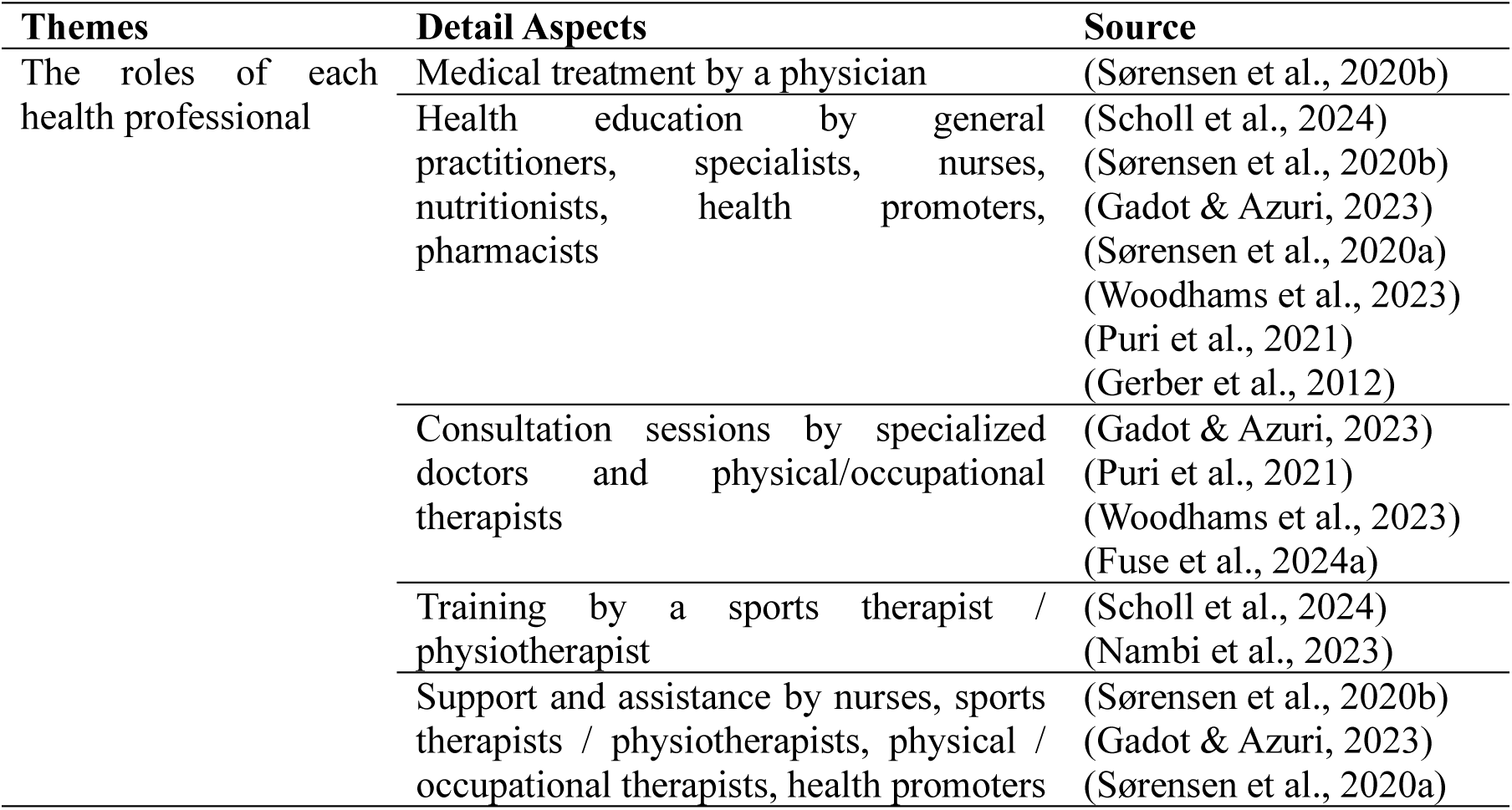
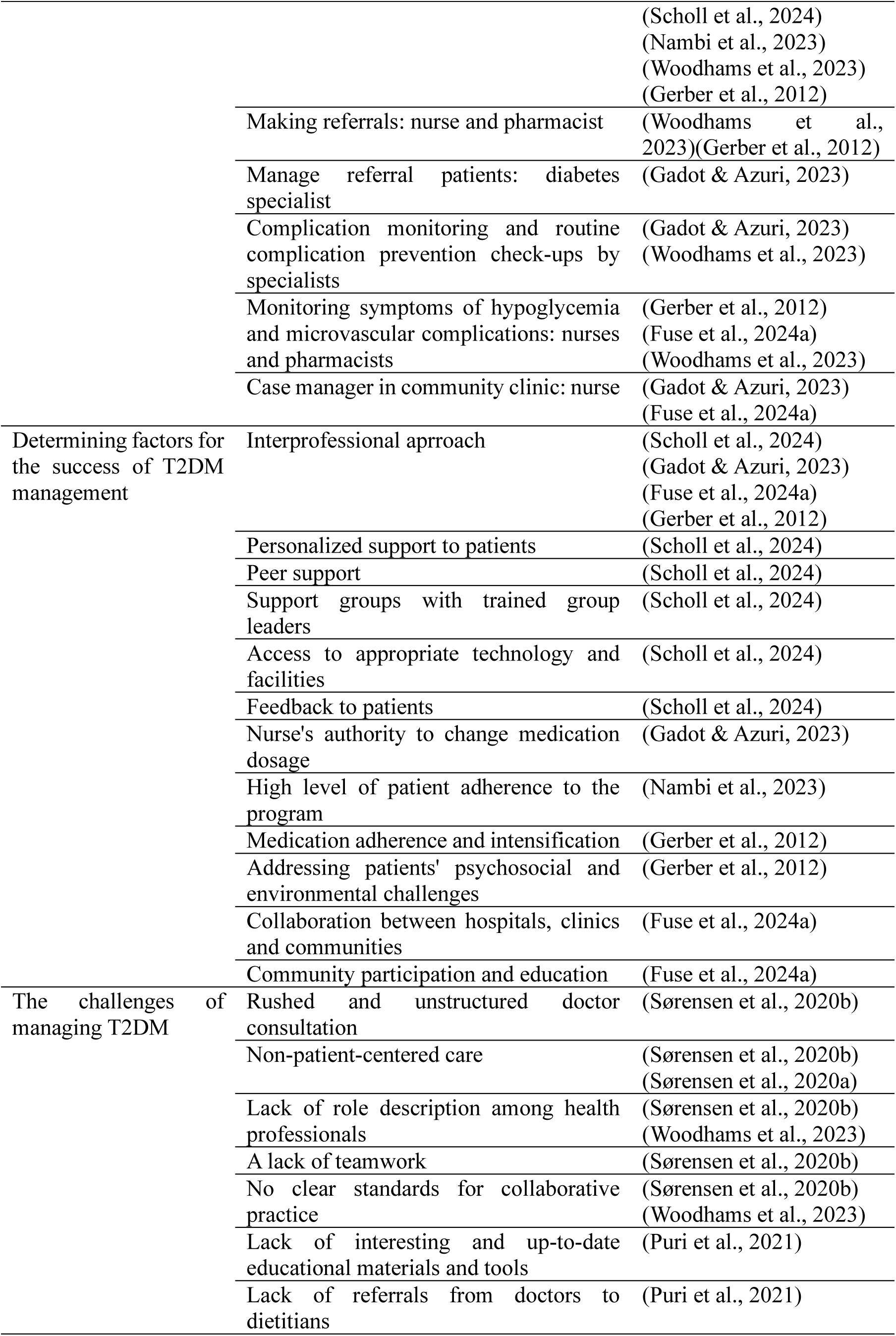
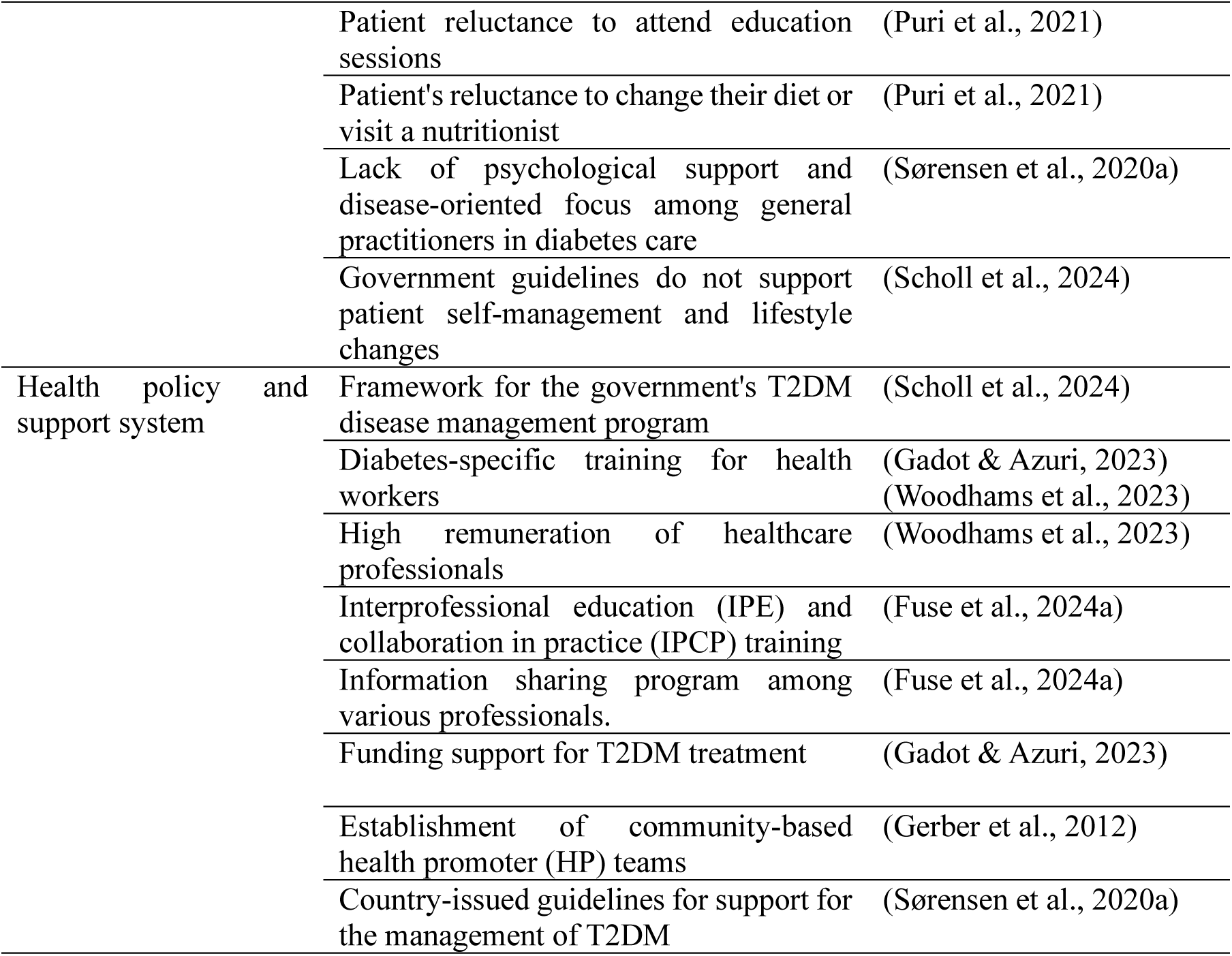
Themes Identified.

### The role of each health professional

Several studies indicate a common and clear role for each professional in the management of people with T2DM in the community. Doctors play a role in providing medical care (Sørensen et al., 2020b), specialist doctors receive referrals, carry out routine monitoring and examinations for complications, and provide consultations (Woodhams et al., 2023), nurses carry out diabetes care in a structured and continuous manner (Gadot & Azuri, 2023; Sørensen et al., 2020b, 2020a), pharmacists providing medication services (Gerber et al., 2012), and nutritionists provide education and consultation services related to nutrition (Fuse et al., 2024b; Puri et al., 2021; Scholl et al., 2024; Woodhams et al., 2023).

However, several studies show almost the same role implementation among health professionals. For example, in terms of providing education. Education was provided by nearly all types of health professionals involved in the study. Several studies mention the type/topic of education given to people with T2DM (Gerber et al., 2012; Scholl et al., 2024), but some studies do not specifically mention it (Gadot & Azuri, 2023; Gerber et al., 2012; Sørensen et al., 2020b, 2020a). Education regarding medication is provided by doctors (Scholl et al., 2024) and pharmacists (Gerber et al., 2012), while nutritionists provide nutritional education and counseling (Fuse et al., 2024b; Puri et al., 2021; Scholl et al., 2024; Woodhams et al., 2023). The study from (Fuse et al., 2024b) provided the same role for nurses and pharmacists in monitoring symptoms of hypoglycemia.

Several health professionals are involved in providing support. Nurses provide emotional support and self-management (Gadot & Azuri, 2023; Sørensen et al., 2020b, 2020a), physical therapists and exercise physiologists provide support and assistance in patients’ physical activities (Fuse et al., 2024b; Woodhams et al., 2023), pharmacists provide support for self-care behavior related to microvascular complications (Woodhams et al., 2023), nutritionists and health promoters provide support for changes in patient behavior/healthy lifestyle (Puri et al., 2021; Scholl et al., 2024; Woodhams et al., 2023), and clinical psychology provides motivational support (Scholl et al., 2024). Sports therapists and physiotherapists even provide group training and supervision for special groups for remote self-managed training sessions (tele-physical therapy) (Nambi et al., 2023; Scholl et al., 2024).

In the study from Scholl et al. (2024), doctors play a role in forming peer support groups, selecting leaders, sending and discussing feedback reports to patients. Meanwhile, in a study from (Gadot & Azuri, 2023), specialist nurses acted as diabetes case managers in community clinics. Consultation sessions are carried out by several health professionals, including specialist doctors who provide consultations regarding complications of T2DM (Gadot & Azuri, 2023; Woodhams et al., 2023), physical/occupational therapists providing consultations regarding the patient’s walking and exercise habits (Fuse et al., 2024b), while nutritionists provide nutrition-related counseling (Fuse et al., 2024b; Puri et al., 2021).

### Determining factors for the success of T2DM treatment

Various factors determine the success of T2DM treatment in this study. Factors can come from health professionals, patients, and the type of intervention provided. The factor most frequently mentioned in studies as determining the success of T2DM treatment is the interprofessional collaboration (IPC) approach (Fuse et al., 2024b; Gadot & Azuri, 2023; Gerber et al., 2012; Scholl et al., 2024). IPC is an approach to treating T2DM that involves the collaboration of several health professionals, including general practitioners together with sports therapists, nutritionists and psychologists (Scholl et al., 2024), specialist nurses together with specialist doctors (Gadot & Azuri, 2023), nurses together with pharmacists, nutritionists, laboratory technicians, physiotherapists, occupational therapists and general practitioners (Fuse et al., 2024b), and pharmacists together health promoter (Gerber et al., 2012).

The IPC model developed in the research of (Gerber et al., 2012) is a collaboration between pharmacists and health promoters. The pharmacist’s role is to provide comprehensive treatment and disease management services (needs assessment, goal setting, patient education, and primary care to intensify therapy as needed), while the health promoter will make home visits and telephone calls to provide education, evaluate medication use, encourage change behavior, and reinforce pharmacist recommendations. Health promoters build close collaborations with pharmacists to support medication management and adherence.

In Scholl et al.’s research, a collaborative model was developed in the form of a personalized self-management program (PSG), where general practitioners are responsible for registering patients and selecting PSG leaders based on communication and organizational skills. Sports therapists lead group-based training sessions, and experts from various professions provide digital education classes (DEE) and telephone coaches with backgrounds in psychology, nutrition, and/or sports science to conduct the telephone training (TC) component (Scholl et al., 2024).

Fuse et al. (2024b) developed a collaborative model involving nurses, pharmacists, nutritionists, physical/occupational therapists, dentists, and health fitness programmers. Nurses and pharmacists play a role in monitoring symptoms of hypoglycemia and contacting patients who do not attend appointments. Dietitians play the role of asking about patient diets, physical/occupational therapists ask about patients’ walking and exercise habits, care managers play a role in hospital "open rounds" to share information and skills. Public health nurses, dentists and health fitness programmers are also involved in the IPC team but their specific roles are not mentioned. In research of Gadot & Azuri (2023), it involved collaboration between specialist nurses and specialist doctors. Diabetes clinic specialist nurses are responsible for managing the care of people with uncontrolled diabetes in community primary care clinics, while specialist diabetes doctors are available part-time to provide additional consultations to patients, if required. Another success factor in terms of service delivery is the presence of feedback to patients provided by doctors (Scholl et al., 2024). The PSG leader will report the patient’s condition to the doctor, then the doctor will follow up and discuss feedback with the patient for better treatment. This study also reported success with appropriate peer group support and trained group leaders. Access to technology by patients and the provision of adequate facilities is one of the supporting factors for successful treatment of T2DM (Scholl et al., 2024). A high level of patient compliance with treatment and the Tele-Physical Therapy (TPT) program provided was reported by (Nambi et al., 2023) and (Gerber et al., 2012) as a determining factor for the success of the intervention.

Gerber et al. (2012) found that one of the things that can influence the success of treatment is overcoming the psychosocial and environmental challenges of people with T2DM. Meanwhile (Fuse et al., 2024) emphasizes the importance of collaboration between hospitals, clinics and communities as well as involving community participation and education in the successful treatment of T2DM.

### Challenges in managing T2DM

Studies show various challenges in the successful treatment of T2DM in the community, especially in the implementation of each role of health professionals. There are studies that find barriers in the form of a lack of clear role descriptions between health professionals and no clear standards for implementing collaborative practice (Sørensen et al., 2020b; Woodhams et al., 2023). There was a lack of teamwork and a lack of joint care planning between general practitioners and other health professionals. GPs focus more on the biomedical aspects rather than the emotional/psychosocial needs of patients (Sørensen et al., 2020b), and time pressures impact the rushed nature of GP consultations, thereby hindering patient engagement and making it difficult to create patient-centred care (Sørensen et al., 2020b, 2020a).

A study found that there were challenges in terms of patient non-compliance with the program. Puri et al., (2021) identified a lack of interesting and up-to-date educational materials and tools and reported patient reluctance to attend nutrition education sessions. In addition, patients were also reported to be reluctant to change their diet or visit a nutritionist for consultation.

### Health policy and support systems

Several studies reported the existence of health policies and support systems in treating T2DM in the community. The study from Scholl et al. (2024) in Germany expressed support from the government through disease management programs (DMPs), which became the basis for the P-SUP intervention in their study. DMPs provide a framework for chronic disease management, including routine check-ups and coordination between various health care providers. In Norway, the government developed guidelines for managing MDT2 to include self-management support and personalized care (Sørensen et al., 2020a).

In Israel, support is provided from the Ministry of Health by authorizing registered nurses (RNs) with special training in diabetes to adjust patient medication doses within certain limits (Gadot & Azuri, 2023), while in Australia, the government provides appropriate training and remuneration. it is appropriate for pharmacists to provide the proposed screening, monitoring and referral services for people with T2DM. Support for patients is also provided by providing health funds, one of which is to develop a case management model for patients with T2DM in community clinics (Gadot & Azuri, 2023). The study from (Fuse et al., 2024b) mentioned policy support from the government through the establishment of the Uonuma School for Public Health and Social Care in 2011 to provide interprofessional education (IPE) and collaboration in practice (IPCP) training for medical students, health and social care professionals, and community members in Japan.

## Discussion

This scoping review seeks to identify and analyze the various roles of health professionals in the management of T2DM, including supporting and inhibiting factors. The role of each health professional in the study varied. Several studies indicate a common and clear role for each professional in the management of people with T2DM in the community. However, several studies show that the implementation of almost the same roles among health professionals, for example providing education and consultation. Some studies mention specific education and consultation topics from each health professional, but some studies do not mention them specifically. Several studies also established a new role in relation to T2DM treatment programs in the community, for example the establishment of a peer support group managed by a doctor, or a specialist nurse who acts as a diabetes case manager in a community clinic.

The unclear role and even overlapping roles of health professionals in treating T2DM can be caused by a lack of clear role definition. This condition can cause confusion about who is responsible for certain tasks, for example insulin initiation and patient education (Manski-Nankervis et al., 2014). There is often uncertainty regarding the competency and capacity of various health professionals, further complicating role clarity. This can result in inefficiencies and gaps in patient care (Manski-Nankervis et al., 2014). A lack of clear role definitions and responsibilities can lead to role ambiguity, which is associated with increased stress, job dissatisfaction, and burnout among healthcare professionals. This ambiguity can result in overlapping or overlooked tasks, compromising the quality of patient care (Cengiz et al., 2021; Kalkman, 2018).

The absence of standardized treatment protocols and clear interprofessional guidelines can also lead to confusion about responsibilities. This often results in duplication of effort or missed responsibilities, which impacts the overall quality of care (Manski-Nankervis et al., 2014). Other causes according to Manski-Nankervis et al. (2014) is fragmented communication. Effective management of T2DM requires good communication and relationships among healthcare professionals. However, varying levels of communication and trust can hinder the coordination necessary for effective diabetes management.

According to Reeves et al. (2017), the cause of not optimal treatment of T2DM is the lack of coordination and collaboration between health professionals involved in the service. Agreeing with this, Riddle (2018) stated that the current service delivery system is still fragmented, lacking clinical information capacity, multiple services and poor design in coordinating the provision of chronic care. Efforts are needed to overcome ambiguity and overlapping roles so that appropriate treatment can be obtained to improve outcomes for T2DM patients in the community.

According to Manski-Nankervis et al. (2014), it is necessary to improve communication and relationships among health professionals so as to help them clarify their roles and responsibilities and understand their specific tasks. Health professionals need to be provided with regular training and education to increase their competence and confidence in managing T2DM, leading to better patient outcomes (Ameh et al., 2024; Manski-Nankervis et al., 2014; Torres et al., 2010). Manski-Nankervis et al., (2014) emphasized the need to establish clear and standardized care protocols and guidelines that help delineate responsibilities and reduce overlap in duties among health care professionals.

Management of T2DM requires an approach that is organized, systematic and involves health professionals in accordance with their field of knowledge, and is patient-centered as a top priority (American Diabetes Association, 2016). According to Proctor & Brown (2024), IPC helps in establishing specific competencies and roles for various health professionals. Effective IPC involves structured communication and coordination among health professionals. This ensures that each professional understands their specific responsibilities and reduces the risk of role overlap (Vyt, 2008).

In this study, the IPC approach was most frequently mentioned as a determining factor in the success of treating T2DM in the community. Effective IPC involves a team of healthcare providers, including physicians, pharmacists, nutritionists, and diabetes educators, working together to manage patient care (Gucciardi et al., 2016). Several previous studies have proven that treating T2DM through an IPC approach significantly improves patient outcomes, including better glycemic control (Champion et al., 2024; Madsen et al., 2022), increased patient satisfaction through the holistic and coordinated care they received (Nurchis et al., 2022), improved self-care and quality of life (Nurchis et al., 2022), and can reduce the economic burden of diabetes management by reducing direct medical costs and increasing medication adherence (Lum et al., 2024).

The implementation of IPC in several studies has experienced several obstacles, namely the lack of clear descriptions of roles between health professionals and the lack of clear standards for implementing collaborative practice (Sørensen et al., 2020b; Woodhams et al., 2023). (Sørensen et al. (2020b) found a lack of teamwork and joint care planning between general practitioners and other health professionals. GPs focus more on the biomedical aspects rather than the emotional/psychosocial needs of patients (Sørensen et al., 2020b), and time pressures impact the rushed nature of GP consultations, thereby hindering patient engagement and making it difficult to create patient-centred care (Sørensen et al., 2020b, 2020a).

According to Gucciardi et al. (2016); Layani et al. (2023), it is necessary to clarify roles and responsibilities through formal agreements and regular team meetings to increase understanding and collaboration among team members. Team members need to implement structured communication models, such as regular case discussion conferences to improve coordination and optimize diabetes management (Gucciardi et al., 2016; Li et al., 2024). It is important to provide continuing professional education and interprofessional education courses to increase healthcare providers’ competency and confidence in collaborative care (Kangas et al., 2021).

Support from policymakers is needed in several ways, such as supporting the financial aspects of IPC and investing in information technology (IT) systems that facilitate seamless information sharing among health service providers (Bawab et al., 2023; Nurchis et al., 2022). Policymakers should promote IPC and provide the necessary resources and infrastructure to support collaborative practice (McGill et al., 2017; Nurchis et al., 2022). Policies need to address the financial and structural barriers that hinder IPC, such as inadequate resources, lack of role clarity, and power dynamics among health care professionals (Bawab et al., 2023; Desse et al., 2023). There is a need to provide additional support to small community health centers so that they can offer IPC despite limited resources (Miller-Rosales & Rodriguez, 2021).

Other studies report several success factors such as the formation of peer support groups with trained group leaders (Scholl et al., 2024) and treatment services to address psychosocial and environmental challenges (Gerber et al., 2012). This shows the importance of providing comprehensive care to people with T2DM. Involving patients as active participants in their care through education and self-management support can lead to better outcomes. This includes providing patient education by a variety of healthcare professionals and involving patients in the decision-making process (McGill et al., 2017; Torti et al., 2022).

In this study, access to technology by patients and the provision of adequate facilities were reported as one of the determining factors for successful treatment of T2DM (Scholl et al., 2024). Several previous studies have found many benefits in using technology for treating T2DM patients in the community. Technologies such as mobile apps, continuous glucose monitors, and smart pens help patients for effective self-management (Grant & Golden, 2019; Kelly et al., 2018). Telemedicine and e-health platforms increase access to healthcare providers and diabetes educators, especially for those in remote or underserved areas resulting in better clinical outcomes and patient well-being (Eberle & Stichling, 2021; Grant & Golden, 2019; Mitchell et al., 2023). Technology-based psychosocial interventions have been shown to reduce diabetes distress and increase self-efficacy, which is critical for managing the emotional aspects of living with T2DM (Bassi et al., 2021; Yap et al., 2024).

Along the way, the use of technology in treating T2DM in the community has faced many challenges. According to (Agarwal et al., 2022; Grant & Golden, 2019), the high costs of advanced diabetes technologies and lack of insurance coverage can be significant barriers to widespread adoption. There are disparities in diabetes technology use among various racial, ethnic, and socioeconomic groups, which may exacerbate or widen health disparities (Agarwal et al., 2022; Jain et al., 2020). Other obstacles include the patient’s personal attributes such as poor competence with technology, low literacy, and language barriers (Jain et al., 2020). Patients often do not receive adequate support or encouragement from healthcare professionals to use diabetes management technologies (Bults et al., 2022; Jeffrey et al., 2019).

Several solutions to obstacles to the use of technology were presented by several previous researchers. According to Bults et al. (2022) and (Jeffrey et al., 2019), it is important to train health care providers to better support and encourage the use of technology in diabetes management. To increase patient engagement and compliance, it is necessary to develop culturally and linguistically adapted interventions (Blasco-Blasco et al., 2020; Lim et al., 2021). Easy-to-use technological designs can be developed through applications with intuitive navigation and visual representation of health data to increase usability (Jeffrey et al., 2019; Yu et al., 2023). Government support through insurance coverage for diabetes management technology can ease the financial burden (Bults et al., 2022; Jeffrey et al., 2019).

### Implications

Health professionals play an important role in managing type 2 diabetes mellitus (T2DM) in the community. Their responsibilities include a variety of activities aimed at improving patient outcomes through education, support, and direct care (Egbujie et al., 2018). This review found a lack of clarity and overlapping roles between health professionals, while people with T2DM need comprehensive and coordinated care. Several studies have provided treatment to patients using an IPC approach, but have not involved all the health professionals needed to treat people with T2DM in the community. Barriers to IPC include unclear roles, lack of teamwork, a lack of IPC models that can be adopted, unclear collaboration pathways, and a lack of training related to IPC. Several studies report successful treatment through access to technology by patients and the provision of adequate facilities. This has been shown to improve outcomes for people with T2DM. However, several studies mention challenges in the form of large costs required to access services, poor patient competency with technology, and even lack of support from health professionals in its use.

This study highlights the importance of support to health professionals from policy makers in the form of regular training and education to increase their competence and confidence in managing T2DM, thereby leading to better patient outcomes (Ameh et al., 2024; Torres et al., 2010). Additionally, it is necessary to develop clear, standardized care protocols and guidelines to help delineate responsibilities and reduce overlap in duties among health care professionals, including models of interprofessional collaboration (Manski-Nankervis et al., 2014). Policymakers need to promote IPC and provide the necessary resources and infrastructure to support collaborative practices with technology integration (McBain et al., 2018).

Further research is needed to establish a clear role for each health professional in the management of people with T2DM in the community. It is also necessary to develop an interprofessional collaboration model so that health professionals can provide comprehensive and coordinated treatment to patients. This includes workflows and communication protocols among health professionals. Furthermore, it is necessary to conduct research by integrating technology in interprofessional collaboration so that health professionals can build more effective communication and provide more affordable services to produce better outcomes.

### Limitations

This scoping review includes only published studies, and only in English. This review may not include every publication related to the role of health professionals in managing T2DM patients in the community. We did not conduct a meta-analysis of quantitative studies that might have identified role characteristics of individual professionals or interprofessional collaborations that are more effective in treating T2DM patients in the community more appropriately than those most commonly listed.

## Conclusion

Some studies reported a general and clear role of each professional in the care of people with T2DM in the community, but some studies mention the implementation of almost the same roles among health professionals. This suggests a lack of clear role definitions and standardized protocols among healthcare professionals managing T2DM resulting in overlapping responsibilities and inefficiencies in patient care. Several studies have used an IPC approach but have not involved all the health professionals needed to treat people with T2DM in the community. This requires policies from governments to promote IPC and provide the necessary resources and infrastructure to support collaborative practices. An IPC model is needed that integrates technology to facilitate communication between health professionals and provide more affordable services resulting in better outcomes.

## Data Availability

All data produced in the present work are contained in the manuscript

